# Distinct mutational profile of Lynch syndrome colorectal cancers diagnosed under regular colonoscopy surveillance

**DOI:** 10.1101/2020.08.21.20179127

**Authors:** Aysel Ahadova, Pauline L. Pfuderer, Maarit Ahtiainen, Alexej Ballhausen, Lena Bohaumilitzky, Svenja Kösegi, Nico Müller, Yee Lin Tang, Kosima Kosmalla, Johannes Witt, Volker Endris, Albrecht Stenzinger, Magnus von Knebel Doeberitz, Hendrik Bläker, Laura Renkonen-Sinisalo, Anna Lepistö, Jan Böhm, Jukka-Pekka Mecklin, Toni T. Seppälä, Matthias Kloor

## Abstract

**Background:** Regular colonoscopy even with short intervals does not prevent all colorectal cancers (CRC) in Lynch syndrome (LS). In the present study, we asked whether cancers detected under regular colonoscopy surveillance (incident cancers) are phenotypically different from cancers detected at first colonoscopy (prevalent cancers).

**Methods:** We analyzed clinical, histological, immunological and mutational characteristics, including panel sequencing and high through-put coding microsatellite (cMS) analysis, in 28 incident and 67 prevalent LS CRCs.

**Results:** Incident cancers presented with lower UICC and T stage compared to prevalent cancers (p< 0.0005). The majority of incident cancers (21/28) were detected after previous colonoscopy without any pathological findings. On the molecular level, incident cancers presented with a significantly lower *KRAS* codon 12/13 (1/23, 4.3% vs. 11/21, 52%; p = 0.0005) and pathogenic *TP53* mutation frequency (0/17, 0% vs. 7/21, 33.3%; p = 0.0108,) compared to prevalent cancers; 10/17 (58.8%) incident cancers harbored one or more truncating *APC* mutations, all showing mutational signatures of mismatch repair (MMR) deficiency. The proportion of MMR deficiency-related mutational events was significantly higher in incident compared to prevalent CRC (p = 0.018).

**Conclusions:** LS CRC diagnosed under regular colonoscopy surveillance are biologically distinct, suggesting that the preventive effectiveness of colonoscopy in LS depends on the molecular subtypes of tumors.

## INTRODUCTION

Individuals with Lynch syndrome (LS), the most common hereditary colorectal cancer (CRC) syndrome, have a 50% lifetime risk of developing CRC^1^. LS is caused by pathogenic variants in one of the Mismatch Repair (MMR) genes *MLH1, MSH2, MSH6*, or *PMS2*^2^.

Due to loss of MMR function, base mismatches occurring during DNA replication remain uncorrected and lead to insertion/deletion mutations (indels), particularly at repetitive sequences (microsatellites). Thus, cancers arising in LS exhibit the molecular phenotype of microsatellite instability (MSI). When indel mutations hit coding microsatellites (cMS), two possible biologically relevant consequences follow: first, mutations at cMS can lead to inactivation of tumor suppressor genes, contributing to carcinogenesis^3^; second, such mutations shift the reading frame and lead to generation of frameshift peptides (FSP), rendering MSI tumors highly immunogenic^4 5–8^.

Surveillance by colonoscopy is a recommended preventive measure in LS mutation carriers^9,10^. Colonoscopy has been shown to decrease the CRC incidence and mortality^11–14^. However, in contrast to the general population^15–17^, a substantial proportion of LS mutation carriers develop “incident carcinomas”, or “post-colonoscopy CRC”^11,18–24^ despite regular colonoscopy. In fact, recent prospective studies^22,23,25^ collecting evidence from patients under surveillance demonstrated no difference in cumulative cancer incidence up to the age of 70 years when compared to studies on retrospective cohorts without surveillance^26–28^.

In parallel to technical, colonoscopy quality-related explanations for the high incidence of CRC under surveillance in LS, biological explanations have been proposed, suggesting that incident cancers may develop from a precursor lesion more difficult to detect than conventional adenomas, such as MMR-deficient crypts^29–32^. MMR-deficient crypts are morphologically undistinguishable from normal colonic crypts, but they lack the MMR protein expression on the molecular level^32–34^. Like MSI CRC, MMR-deficient crypts also present with MSI and MSI-induced tumor suprressor gene mutations as a direct consequence of MSI, thus possessing the theoretical potential to develop into cancer. However, direct evidence of such a progression is not trivial to obtain, as no technical means to monitor MMR-deficient crypts exist.

In contrast to clinical characteristics^35^, the molecular properties of incident cancers have not been characterized so far. We aimed to analyze the molecular characteristics of incident LS CRCs diagnosed under regular surveillance and to compare them with prevalent LS CRCs diagnosed at first colonoscopy or prior to surveillance.

## METHODS

### Patients and tumor samples

Carriers of pathogenic MMR variants that underwent colonoscopy surveillance with a planned 3-year interval (2 years if previous CRC) were identified from the prospectively maintained Finnish Lynch syndrome registry. Available formalin-fixed paraffin-embedded (FFPE) tumor blocks from patients who developed incident (n = 28) and prevalent (n = 7) cancers were collected from the Lynch Syndrome Biobank at the Central Finland Central Hospital, Jyväskylä, Finland. FFPE tumor tissue blocks from LS patients with prevalent CRC (n = 60) were collected at the Department of Applied Tumour Biology, Institute of Pathology, University Hospital Heidelberg as part of the German HNPCC Consortium. All patients provided an informed and written consent, and the study was approved by the Institutional Ethics Committee. The DNeasy FFPE Kit was used for the isolation of tumor DNA after manual microdissection from 5–6 µm thick hematoxylin/eosin (HE)-stained FFPE tissue sections (Qiagen, Germany).

### Mutation analysis

Targeted next generation sequencing was performed as described previously on IonTorrent S5XL/Prime sequencer using a custom 180 amplicon panel (CRC panel) encompassing mutation HotSpot regions in 30 genes^36,37,38,39^. Data analysis was performed using the Ion Torrent Suite Software (version 5.10). Only variants with an allele frequency > 5% and minimum coverage > 100 reads were taken into account. Variant annotation was performed using Annovar (hg19 genome)^38^. Annotations included information about nucleotide and amino acid changes of RefSeq annotated genes, COSMIC and dbSNP entries as well as detection of possible splice site mutations. For data interpretation and verification, the aligned reads were visualised using the IGV browser (Broad Institute)^39^.

MSI analysis was performed using a sensitive and specific mononucleotide marker panel (BAT25, BAT26, and CAT25) as described previously^40^. cMS mutation analysis was performed using a novel high-throughput method for quantitative fragment length analysis with 5-carboxyfluorescein-labeled primers specific for a set of 22 cMS^41^, (see Supplementary Table 1 for details), which were selected based on two criteria: evidence of a functional driver role of mutation^41^ and potential significance as a source of immunogenic frameshift peptide neoantigens^42^. PCR products were visualized on an ABI3130xl sequencer, and the obtained results were processed using the ReFrame algorithm to obtain quantitative estimation of the frequency of the mutant alleles in tumor specimens^43^. Mutation status of *B2M* was determined by Sanger sequencing as described previously^44^. The obtained mutational data for incident cancers were compared with the mutational data for prevalent cancers published in our previous reports^31,36,43^.

**Table 1:** Clinical characteristics of patients with incident CRC. M – male, F – female, LG – low grade, CUP – carcinoma with unknown primary. *-Finding at last colonoscopy column refers to the previous colonoscopy before cancer diagnosis.

| Patient | Age at diagnosis | Gender | Location | TNM Stage | UICC Stage | Gene | Age at last FU | Age at death | Cause of death | Months since last colonoscopy | Reason of examination | Finding at last colonoscopy* |
| --- | --- | --- | --- | --- | --- | --- | --- | --- | --- | --- | --- | --- |
| 1 | 54.6 | M | splenic flexure | T2N0M0 | I | MLH1 | 62.9 |  |  | 21.0 | symptoms | 0 |
| 2 | 61.7 | F | ascendens | T1N0M0 | I | MLH1 | 76.5 |  |  | 20.3 | follow-up | Advanced lesion |
| 3 | 44.2 | M | descendens | Dukes A | I | MLH1 | 55.1 |  |  | 36.0 | follow-up | LG adenoma |
| 4 | 69.7 | F | sigmoid | T1N0M0 | I | MSH2 | 74.9 | 75.1 | cardiac insufficiency | 7.3 | follow-up | Advanced lesion |
| 5 | 63.1 | F | tranverse | T3N0M0 | II | MSH2 | 72.6 | 72.6 | pancreatic cancer | 25.0 | follow-up | 0 |
| 6 | 70.5 | M | sigmoid | TisNxM0 | 0 | MLH1 | 78.4 |  |  | 24.5 | follow-up | 0 |
| 7 | 35.5 | M | caecum | T3N1M0 | III | MLH1 | 44.3 | 44.3 | gastric cancer | 28.0 | symptoms | 0 |
| 8 | 57.3 | F | sigmoid | T3N0M0 | II | MLH1 | 63.3 |  |  | 30.0 | follow-up | 0 |
| 9 | 54.5 | F | caecum | T2N0M0 | I | MLH1 | 63.4 | 63.4 | CRC | 31.2 | follow-up | 0 |
| 10 | 71.6 | F | rectum | T2N0M0 | I | MLH1 | 75.1 | 75.1 | biliary tract cancer | 24.0 | follow-up | 0 |
| 11 | 41.7 | F | descendens | T3N2M0 | III | MLH1 | 44.7 | 44.7 | CRC | 23.0 | symptoms | 0 |
| 12 | 43.6 | F | ascendens | T1N0M0 | I | MLH1 | 57.5 | 57.5 | breast cancer | 26.0 | follow-up | 0 |
| 13 | 41.7 | F | tranverse | T1N0M0 | I | MLH1 | 50.1 |  |  | 26.0 | follow-up | LG adenoma |
| 14 | 42.4 | F | tranverse | T3N0M0 | II | MLH1 | 48.8 | 48.8 | pancreatic cancer | 24.4 | follow-up | 0 |
| 15 | 71.5 | M | ascendens | T2N0M0 | I | MSH2 | 84.1 |  |  | 36.0 | follow-up | LG adenoma |
| 16 | 43.6 | M | tranverse | T3N0M0 | II | MLH1 | 53.9 | 53.9 | another CRC | 26.0 | follow-up | 0 |
| 17 | 71.9 | F | caecum | T2N0M0 | I | MLH1 | 81.4 | 81.4 | pneumonia | 29.0 | follow-up | 0 |
| 18 | 69.0 | F | caecum | T2N0M0 | I | MSH2 | 69.0 | 69.0 | postoperative complication | 37.7 | symptoms | 0 |
| 19 | 42.0 | F | caecum | T1N0M0 | I | MLH1 | 58.2 |  |  | 39.5 | follow-up | 0 |
| 20 | 35.1 | M | caecum | Dukes B | II | MLH1 | 64.4 |  |  | 37.6 | follow-up | 0 |
| 21.a | 54.2 | F | ascendens | T2N0M0 | I | MLH1 | 63.3 |  |  | 30.1 | follow-up | 0 |
| 21.b | 56.8 | F | sigmoid | T3N0M0 | II | MLH1 | 63.3 |  |  | 28.8 | follow-up | 0 |
| 22 | 54.2 | M | ascendens | T3N0M0 | II | MLH1 | 63.4 | 65.0 | CUP (brain, lung) | 18.0 | follow-up | LG adenoma |
| 23 | 53.8 | M | ascendens | T2N0M0 | I | MLH1 | 58.8 |  |  | 16.4 | follow-up | LG adenoma |
| 24 | 82.8 | M | descendens | T2N0M0 | I | MLH1 | 85.1 |  |  | 24.6 | follow-up | 0 |
| 25 | 55.0 | F | caecum | T1N0M0 | I | MLH1 | 66.4 |  |  | 39.1 | follow-up | 0 |
| 26 | 43.5 | M | caecum | T2N0M0 | I | MLH1 | 52.5 |  |  | 36.2 | follow-up | 0 |
| 27 | 27.2 | M | caecum | T2N0M0 | I | MLH1 | 42.9 |  |  | 37.9 | follow-up | 0 |

### Immunohistochemical staining and quantification of T cell density

FFPE tissue sections (2–3 µm) were used for immunohistochemical staining^45,46^. Briefly, sections were deparaffinized and rehydrated and subsequently stained according to standard protocols. Following primary antibodies were used: anti-CD3 (clone PS1, dilution 1:100, Abcam, Germany); anti-MLH1 (clone G168–15, dilution 1:300, BD Pharmingen, Germany); anti-MSH2 (clone FE11, dilution 1:100, Calbiochem, Germany). As a secondary antibody, the biotinylated anti-mouse/anti-rabbit antibody (Vector Laboratories, was used at 1:100 dilution. Staining was visualized using the Vectastatin elite ABC detection system (Vector Laboratories, Burlingame, CA, USA) and 3,3’-Diaminobenzidine (Dako North America Inc., Carpinteria, CA, USA) as a chromogen. For counterstaining hematoxylin was used. Stained sections were scanned using NanoZoomer S210 slide scanner (Hamamatsu) and viewed using NDP.view2 Viewing Software (Hamamatsu). Four random 0.25 mm^2^ square regions were drawn in the tumor tissue and positive cells in each region were counted using the QuPath Software, giving mean values per 0.25 mm^2^.

### Statistical calculations

Statistical significance of differences between binary variables was calculated using Pearson’s Chi-squared test or Fisher’s exact test. Statistical significance of differences in mutation frequencies of cMS genes, as well as significance of differences in immune infiltration was calculated using two-sided Wilcoxon Rank Sum test (Mann-Whitney) test. Correction for multiple testing was performed using Benjamini-Hochberg procedure. P values smaller than 0.05 were considered statistically significant. All scripts were written in R^47^, version 3.6.0 using the R Studio environment^48^. All 95% confidence intervals (CI) were calculated with modified Wald method.

## RESULTS

### Clinical characteristics

Clinical data and tumor specimens (n = 28) were collected from 27 LS patients who developed incident CRC during the 2–3-yearly preventive colonoscopy surveillance (23 *MLH1* and 4 *MSH2* carriers, 15 females and 12 males). Sixty-seven tumors from LS patients diagnosed with CRC as prevalent cancers were used as a comparison group. Median age at diagnosis was not significantly different between patients with incident and with prevalent cancer (54.4 vs. 50.0 years, p>0.05). Nineteen out of 28 (68%) incident and 33 out of 49 (67%) prevalent cancers with information on tumor localization were localized in the proximal colon. The clinical parameters of incident cancers are summarized in Table 1.

The median duration of follow-up was 8.9 years (range 0.0–29.3 years). Twelve patients with incident cancers died during follow-up due to different reasons. Three of the 12 deceased patients died due to CRC: One patient (#11) died from a symptomatic CRC that was diagnosed only two years after previous uneventful colonoscopy. Patient #16 died from another, metachronous, CRC that was diagnosed after 6 years of not attending scheduled colonoscopies. Patient #9 developed CRC liver metastases 7 years after the operation of a T2N0 caecum cancer, though no other primary tumor was found (Table 1).

Incident cancers presented with lower UICC stage compared to prevalent cancers (p = 0.0002); the majority of incident cancers were stage I, whereas the majority of prevalent cancers were stage II tumors (Figure 1A). T stage of incident cancers was significantly lower in prevalent cancers (p = 0.00004), and no T4 lesions were identified among incident cancers (Figure 1B).

**Figure 1.**
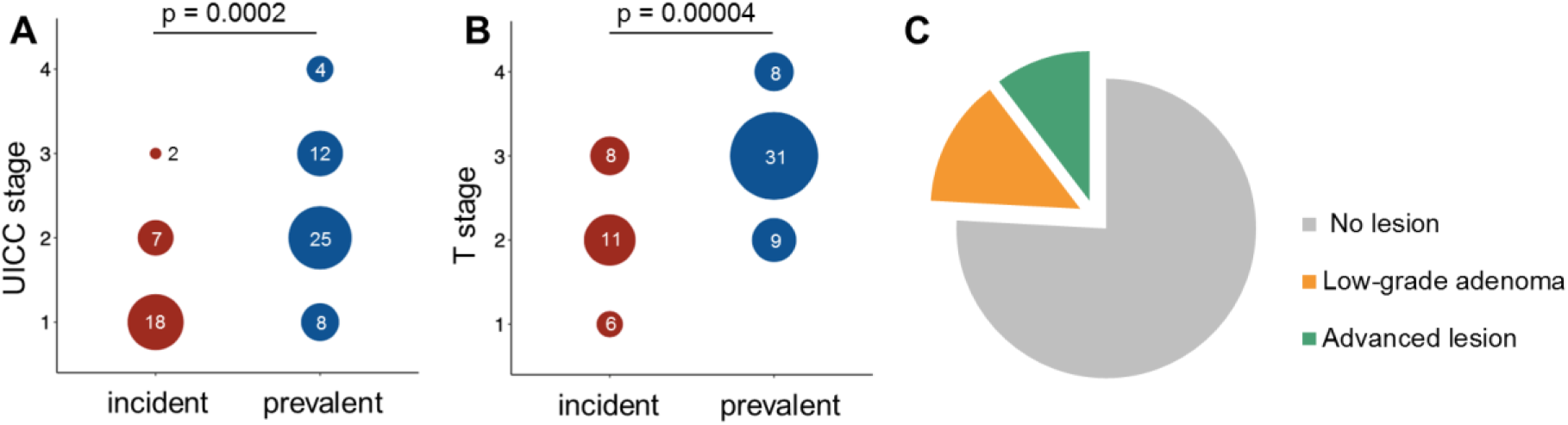
Clinical characteristics of incident cancers. A-B. Distribution of UICC stage among incident and prevalent tumors. Tumors identified under surveillance have significantly lower UICC stage (A) and T stage (B) compared to tumors identified outside of surveillance. C. Findings at previous colonoscopy in patients with incident cancers. The majority of patients with incident cancers did not present with any lesion at previous colonoscopy examination.

The median time since last colonoscopy in patients under surveillance was 27 months (range: 7.3–39.5 months). Time since last colonoscopy did not correlate with the stage of tumor (Supplementary Figure 1). The majority of incident CRCs developed after a colonoscopy in which no lesions were detected (21/28, 75.0%, 95% CI: 56.4–87.6%, Table 1, Figure 1C). All performed colonoscopies were successful and of high quality, with complete examination reaching the remaining colon length and bowel preparation enabling the visualization of the entire mucosal surface.

### Histopathology of incident cancers

Representative HE and immunohistochemistry sections of the incident cancers were examined for microscopic pattern of tumor growth, degree of differentiation and presence of MMR-deficient crypts (Supplementary Table 2).

Among the 22 cases assessable for the tumor growth pattern, 12 showed a sessile (55.5%), 6 (27.3%) showed a pedunculated and 3 (13.6%) showed an undermining growth pattern. All tumors showed at least moderate degree of differentiation, with 16/28 (57.1%) of them exhibiting mucinous components.

MMR-deficient crypts were present in two of the incident cancers: in both cases, these MMR-deficient crypts were present adjacent to areas of high-grade dysplasia/carcinoma *in situ* (Table 1, Patient #6, Figure 2A and Table 1, Patient #22, Figure 2B). The MMR deficient crypt in Patient #22 also showed pronounced immune infiltration (Figure 2B).

**Figure 2.**
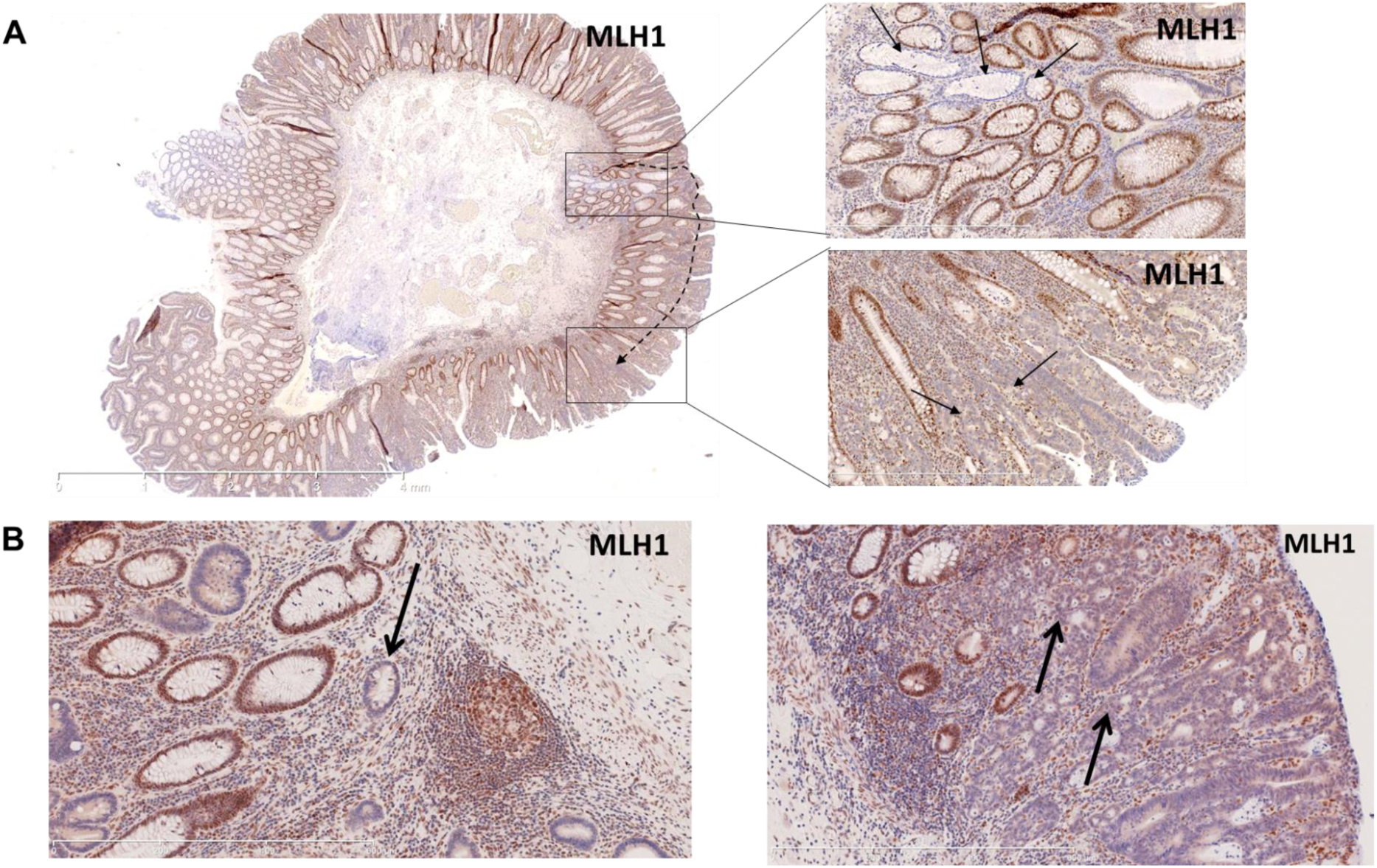
Histology images of tumor specimens with MMR-deficient crypt foci. A. Resection sample with carcinoma in situ arising presumably from an MMR-deficient crypt. On the left panel, the overview of the resected sample (MLH1 staining), on the right upper panel, higher magnification of the MMR-deficient crypt (MLH1 staining), on the right lower panel, higher magnification of carcinoma in situ (MLH1 staining). B. MMR-deficient crypt, MLH1 staining on the left and another region of the same sample showing a non-invasive carcinoma in situ on the right panel.

### Mutational profile and MMR deficiency signatures in incident cancers

We aimed to analyze how MMR deficiency influences mutational events in incident cancers and studied MMR deficiency signatures, namely presence of C>T transitions at CpG sites and insertion/deletion (indel) mutations, in *APC* and *KRAS* mutations observed in incident cancers, and compared to previous sequencing results obtained from prevalent CRC^31,36,49^.

In contrast to the relatively high prevalence of *KRAS* codon 12/13 mutations among prevalent LS CRCs described previously (11/21, 52%^36^), only 1 codon 12 mutation was identified among the analyzed incident tumors (1/23, 4.3%; p = 0.0005) (Figure 3A, Supplementary Tables 3 and 4). Moreover, no pathogenic *TP53* mutations were identified in the analyzed set (0/17), which compared to prevalent cancers (7/21, 33.3%^36^) yielded a significantly lower *TP53* mutation frequency in incident CRCs (p = 0.0108, Figure 3A, Supplementary Tables 3 and 4). The proportion of *CTNNB1-*mutant samples (5/23, 21.7%) in incident cancers was similar to the proportion of *CTNNB1*-mutant tumors detected in prevalent cancers^49^ (10/48, 20.8%; p = 1.0, Figure 3A).

**Figure 3.**
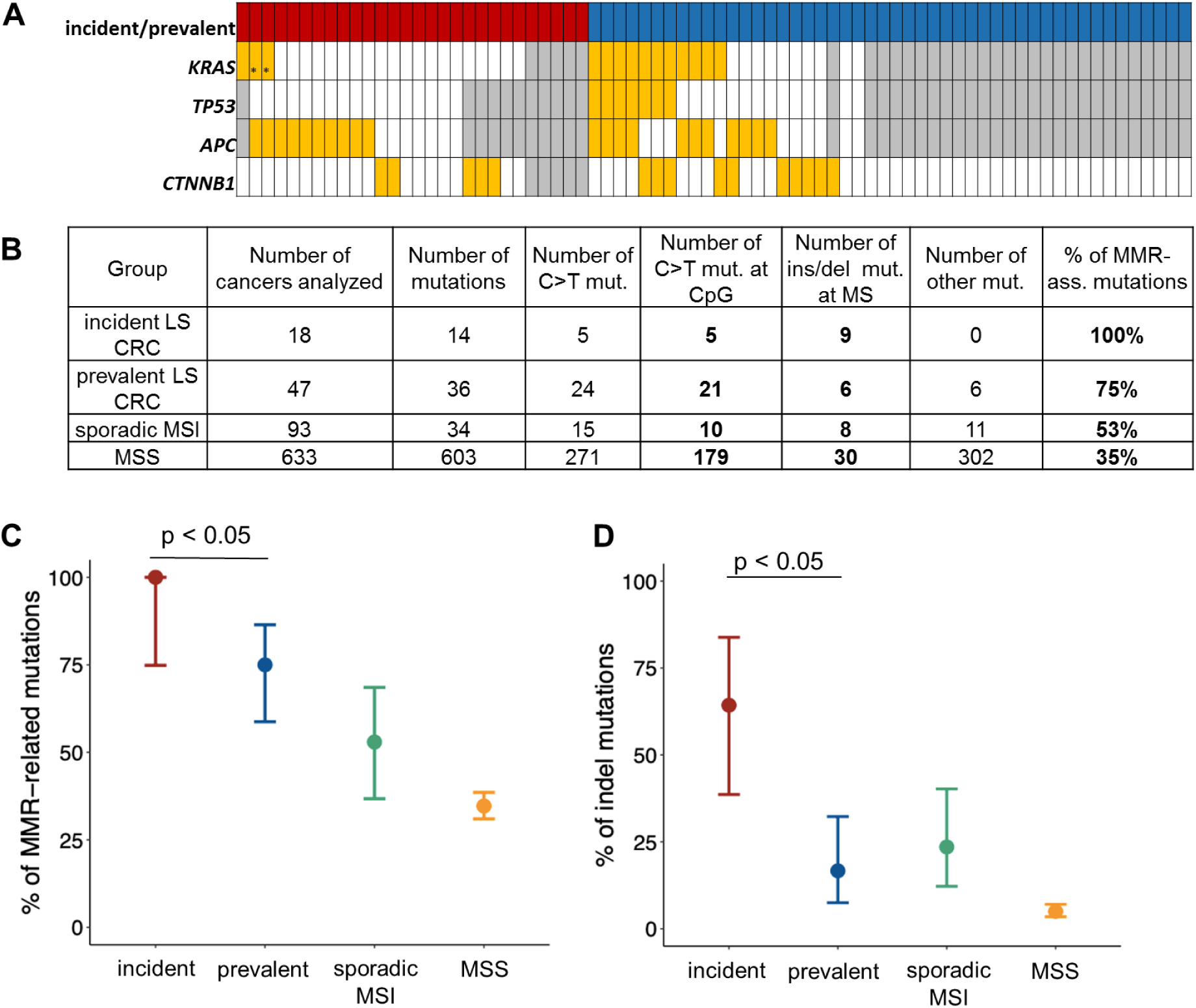
Mutational profile and MMR deficiency signatures in incident LS CRC. A. Mutation status of CRC genes in incident cancers analyzed in this study and prevalent cancers reported before^31,36^ (for cohorts: red – incident CRC, blue – prevalent CRC; for genes: orange – mutant, white – wild type, grey – n.a.; * – KRAS mutations at codons other than codon 12/13). B. Summary of the number of specific MMR deficiency-related mutations in incident LS CRC compared to prevalent LS CRC, sporadic MSI CRC and MSS CRC previously reported in Ahadova et al^36^. C. Comparison of the proportion of all MMR deficiency-related mutations between different CRC groups reveals higher proportion in incident compared to prevalent tumors (100%, 95% CI: 74.85–100% vs. 75%, 95% CI: 58.7–86.4%; Fisher’s exact test, p = 0.0470). D. Comparison of the proportion of indel mutations between different CRC groups reveals higher proportion in incident compared to prevalent tumors(64.3%, 95% CI: 38.6–83.8 vs 16.7%, 95% CI: 7.5–32.3%; Fisher’s exact test, p = 0.0068).

Ten out of 17 incident cancers presented with a total of 14 truncating *APC* mutations (Figure 3A). All 14 detected *APC* mutations represented either C>T transitions at CpG sites or insertion/deletion (indel) mutations, reflecting mutational signatures associated with MMR deficiency and arguing in favor of the early onset of MMR deficiency in LS incident CRC, prior to *APC* mutations. Importantly, the proportion of such mutations was significantly higher in incident cancers compared to prevalent cancers (100% vs 75%, 95% CI: 74.9–100% and 58.7–86.4%, respectively; p = 0.0470, Figure 3B, 3C).

When focusing on indel *APC* mutations alone, a significantly higher proportion of mutations was found in incident CRC compared to prevalent CRC in LS patients (64.3% vs 16.7%, 95% CI: 38.6–83.8% and 7.5–32.3%, respectively; p = 0.0068, Figure 3B, 3D).

### CMS analysis in incident cancers

Mutation frequencies obtained from the quantitative cMS analysis were compared between incident (n = 28) and prevalent (n = 67) tumors across all genes and for each gene. Generally, the frequency of cMS mutations in all 22 analyzed genes was slightly, but significantly elevated in the group of incident cancers compared to prevalent cancers (median 0.35 in incident vs 0.31 in prevalent tumors, p = 0.018, Figure 4A). As mutations at cMS presumably accumulate in association with the progression time of the tumor, we analyzed cMS mutation frequencies in association with the UICC stage. In prevalent LS CRC, we observed a significant increase of the cMS mutation frequencies from UICC I to UICC II (median for UICC I 0.28 vs UICC II 0.36, p = 0.002, Figure 4B), whereas the incident LS CRC presented with high cMS mutation frequencies already in stage I tumors, and no further increase was observed in stage II tumors (Figure 4C). Importantly, the cMS mutation frequency was higher in stage I incident LS CRCs compared to stage I prevalent LS CRCs (median for UICC I in incident tumors 0.35 vs 0.28 in prevalent tumors, p = 0.005, Figure 4D).

**Figure 4.**
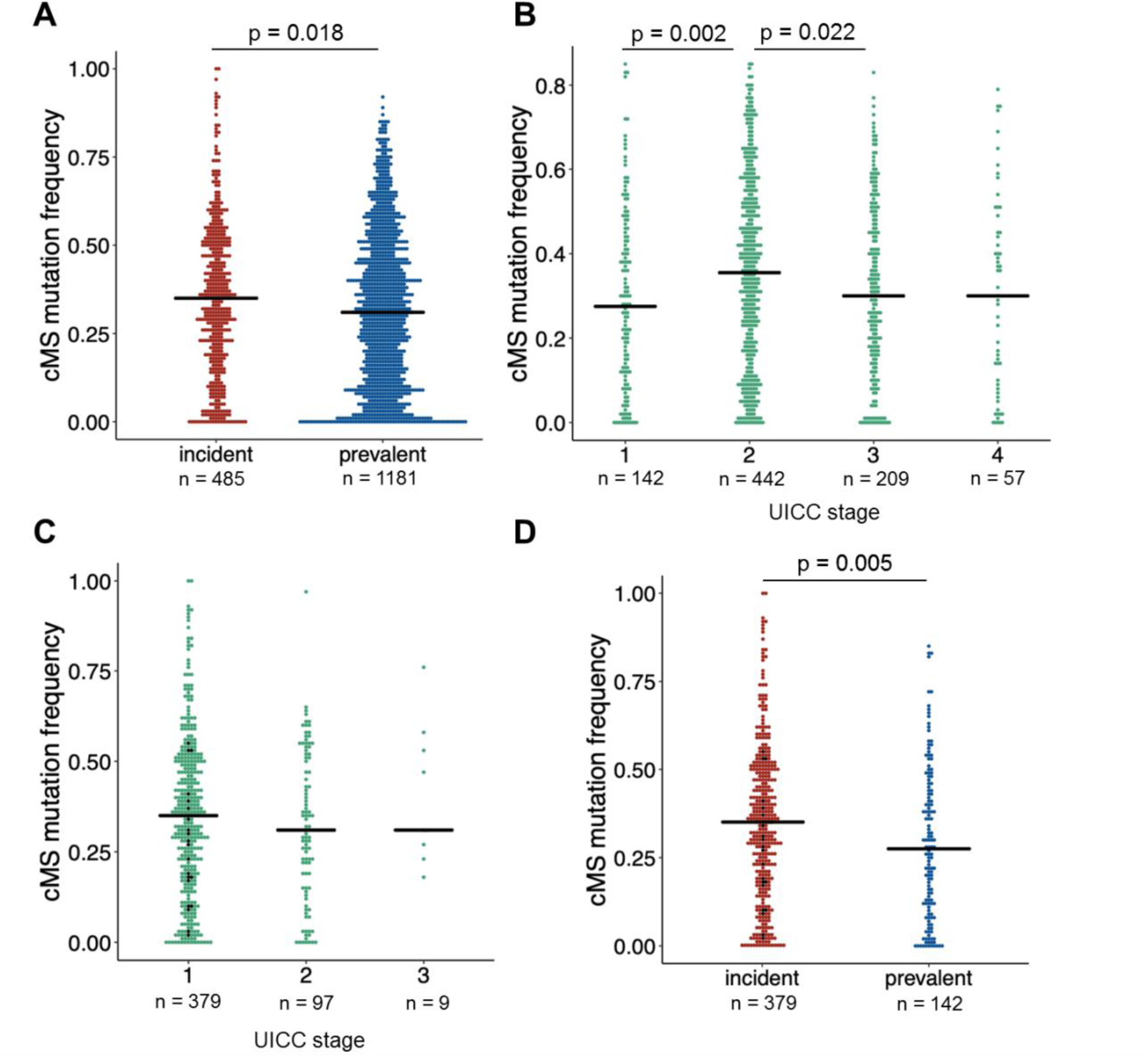
Analysis of coding microsatellite (cMS) mutations in incident and prevalent LS CRC. A. CMS mutation frequency in incident and prevalent LS CRC. B. CMS mutation frequency in prevalent LS CRC by UICC stage. C. CMS mutation frequency in incident LS CRC by UICC stage (stage I group includes data from UICC 0 tumor, see black data points). D. CMS mutation frequency in stage I incident (stage I group includes data from UICC 0 tumor, see black data points) and stage I prevalent LS CRC.

The analysis of mutations in 22 specific cMS genes revealed a significantly higher proportion of mutant alleles in two genes (*LMAN1* (0.29 vs. 0.11) and *ELAVL3* (0.37 vs. 0.17)) and a significantly lower proportion of mutant alleles in one of the analyzed cMS located in the *RFC3* (0.03 vs. 0.19) gene in incident cancers compared to prevalent ones (Supplementary Figure 2 and 3).

### Immune infiltration and immune evasion in incident cancers

We asked whether the early onset of MMR deficiency and the higher proportion of tumors with cMS mutations is reflected by the immune response in incident cancers, and analyzed the CD3-positive T cell infiltration in incident and prevalent LS CRC. As MMR gene-dependent differences of the immunogenicity of LS CRC have been reported before^49,50^, we performed an MMR gene-wise comparison of immune infiltration focusing on the *MLH1*-associated CRCs representing the vast majority in our incident cancer group (24/28). Dense immune infiltration was observed in both incident and prevalent tumor tissue (155 vs. 149 CD3+ cells/0.25mm^2^, respectively) and normal mucosa, although no significant differences between incident and prevalent tumors could be detected (p = 0.6, Figure 5).

**Figure 5.**
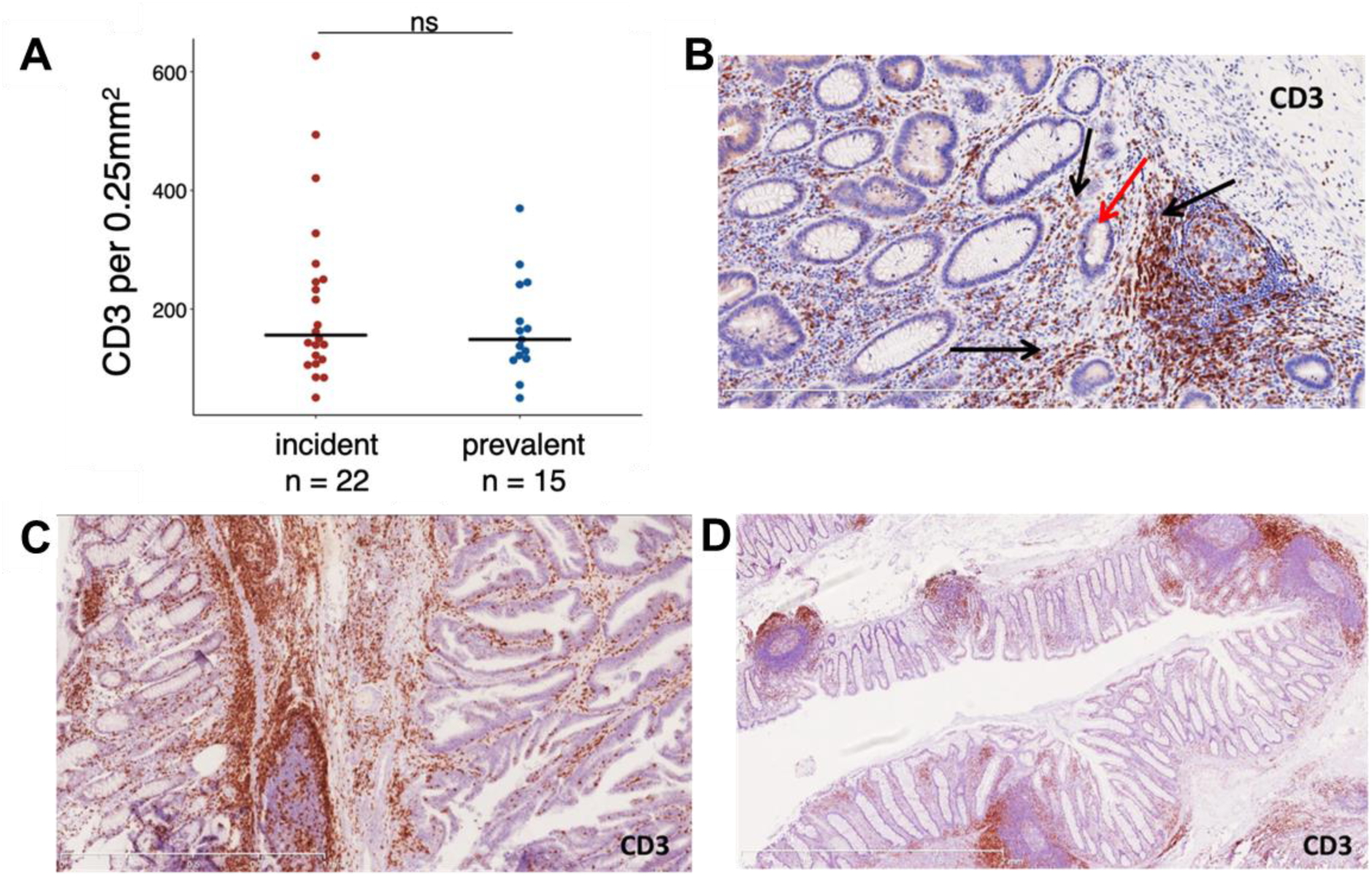
Immune infiltration with CD3-positive T cells in incident and prevalent cancers. A. Immune infiltration in MLH1-associated incident and prevalent LS CRC. B-D. Exemplary CD3 stainings of an MMR-deficient crypt (B, see Figure 2B for the MLH1 staining, red arrow points to the MMR-deficient crypt) tumor (C) and normal mucosa (D) regions of a transverse colon cancer specimen.

The pronounced immune response against MSI CRC often results in the acquisition of *B2M* mutations, the most common mechanism of immune evasion in MSI tumors leading to abrogation of HLA class I-mediated antigen presentation^46,51^. We analyzed *B2M* in incident and prevalent CRCs and found a *B2M* mutation prevalence of 20.8% (5/24) in incident CRC, which was similar to the *B2M* mutation prevalence of prevalent CRC (13/54, 24.1%; p = 1.0).

## DISCUSSION

In the present study, we provide first evidence that incident CRCs in LS are distinct from prevalent cancers with regard to their clinical, histological and mutational characteristics.

Clinically, most incident cancers were of UICC stage I/II and thus of significantly lower stage than the prevalent cancers of our control cohort. Low tumor stage, typical absence of lymph node involvement and a favorable clinical course of incident cancers observed in our study are in line with previous reports^11,18,22,35,52,53^. Only one CRC-related death was clearly associated with a primary CRC included in this study and showing signet ring cell features, associated with poor survival^54^. This mirrors the previously reported excellent survival under prospective observation^55^, which could be attributed to the early detection via colonoscopy.

Histologically, tumors with mucinous components were frequent among incident CRCs in LS. Presentation with mucinous histology in MSI cancer has previously been associated with a high cMS mutational load^56^. The elevated cMS mutation frequency detected in incident cancers of our study (Figure 4) and suggesting the predominance of MMR deficiency-initiated CRC evolution among incident CRCs may therefore be responsible for a high mutational variability resulting in mixed and mucinous histology.

The hypothesis of MMR deficiency as an initiating factor in incident CRC formation is supported by two additional observations: (1) histologically normal MMR-deficient crypt foci were detected in the direct vicinity of two incident CRCs, providing first evidence that MMR-deficient crypts can give rise to incident CRC development in LS; (2) on the molecular level, *APC* mutations in incident CRCs showed a significantly stronger association with signatures of MMR deficiency^57^ than in prevalent CRCs, indicating that MMR deficiency as an early event commonly precedes APC mutations.

Importantly, we found significantly less *KRAS* mutations in incident cancers than previous studies analyzing prevalent CRC in LS^58^. Two scenarios for the observations are possible: (1) colonoscopy with adenoma removal may theoretically be more effective in preventing *KRAS*-mutated lesions, as *KRAS* mutations are associated with conventional adenomas^59,60^. This would imply that incident cancers may develop from other, *KRAS*-wild type lesions that are more difficult to detect. In fact, a recent study analyzing the efficacy of colonoscopy depending on the molecular subtype of tumors in the general population showed a weaker CRC risk reduction after colonoscopy for *KRAS*-wild type tumors^61^. (2) Alternatively, oncogene-activating missense mutations, which need to affect very specific nucleotides and therefore have a lower likelihood per genome replication than indel mutations, may be less frequent in tumors with rapid evolution and short progression times such as incident cancers^62^. This hypothesis could also explain the absence of *TP53* point mutations, which are generally considered late events in colorectal carcinogenesis^63^, and the relative scarcity of *CTNNB1*-activating point mutations in the incident CRC of our study.

In contrast to rare point mutations, the cMS mutation load of incident CRCs was significantly elevated. In addition to the general enrichment, we observed a significantly higher mutation frequency of 2 cMS genes, *LMAN1* and *ELAVL3*, and a significantly lower mutation frequency of the *RFC3* cMS gene. A high proportion of *LMAN1* mutations in LS incident cancers may thus lend support to the previously reported crucial role of cMS mutations during early steps of MSI carcinogenesis^64^.

Colonoscopy quality might be another factor responsible for the development of cancers under surveillance. In our study, colonoscopies performed prior to the examination revealing cancer were documented as complete procedures fulfilling the criteria for a high quality colonoscopy (evidence of full visualization of the remaining bowel length and adequate bowel preparation)^65^. This is in line with the previous observations by Lappalainen et al. showing no association between incident cancers and a prior colonoscopy of compromised quality^66^. Also, the proportion of tumors located in the proximal colon, a localization often associated with lower colonoscopy sensitivity^15,61^, was identical between incident and prevalent tumors analyzed in our study, indicating that localization-related colonoscopy sensitivity alone also does not explain the occurrence of incident CRCs in LS carriers under surveillance. The adenoma detection rate (ADR) in the contributing centers for follow-up colonoscopies has also been shown to be comparable with the previous reports of recent large prospective studies^22,66^.

In line with previous observations reported by the Prospective Lynch Syndrome Database (PLSD)^52^ and other large studies^22,35^, no correlation was observed between time since last colonoscopy and tumor stage among incident cancers. Previous studies reported incident cancer development in the same segment of colon, where previously a polypectomy was performed, in 20–50% of cases^35,67^. Although no information on the localization of a lesion detected at previous colonoscopy was available in this study, adenoma at previous colonoscopy was found in 25% of patients with incident cancers, which is in line with other reports^18,35,66^.

The strength of our study is the first molecular characterization of incident cancers in LS and their comparison to prevalent CRC in LS, as well as high-resolution analysis of MMR deficiency-associated mutational events using a newly established method^43^. The weakness of the study is the analysis of incident cancers from a single country, the majority harboring *MLH1* germline variants and thereby representing only one of the two *MMR* genes most frequently associated with incident cancer^49^. Validation of our results in a larger international multi-center study is therefore warranted in order to include more *MSH2* pathogenic variant carriers to analysis and examine potential differences between *MLH1* and *MSH2*-associated LS, as has been suggested recently^49^.

In conclusion, our study for the first time identifies a set of characteristics that differentiate incident cancers in LS from prevalent cancer occurring without surveillance: a lower tumor stage, a high proportion of tumors with mucinous areas, a predominance of indel mutations over point mutations, and a low prevalence of RAS mutations. This implies that prevention by colonoscopy may shift the molecular manifestation of LS-associated CRCs towards MMR deficiency-initiated cancers highlighting the need for preventive measures targeting MMR-deficient cells directly.

## Data Availability

All data presented in this manuscript are available at the Department of Applied Tumor Biology.

## ADDITIONAL INFORMATION

## Acknowledgements

The excellent technical support of Nina Nelius, Petra Höfler and Lena Ehret-Maßholder is gratefully acknowledged.

## Authors’ contributions

Study concept and design (AA, TS, JPM, MK); acquisition of data (AA, PLP, MA, AB, LB, SK, NM, YLT, KK, JW, VE, AS, LRS, AL, JB, TS); analysis, interpretation of data, manuscript draft (AA, PLP, TS, MK, JPM); critical revision of the manuscript and decision to submit (all authors); obtained funding (TS, JKM, MvKD, HB, MK); study supervision (TS, JKM, MK).

## Ethics approval

All patients provided an informed and written consent, and the study was approved by the Ethics Committee of the University Hospital Heidelberg and Central Finland Hospital District Ethical Committee. The study was performed in accordance with the Declaration of Helsinki.

## Consent for publication

Not applicable

## Data availability

All data presented in this manuscript are available at the Department of Applied Tumor Biology and can be shared upon request.

